# Early phylodynamics analysis of the COVID-19 epidemic in France

**DOI:** 10.1101/2020.06.03.20119925

**Authors:** Gonché Danesh, Baptiste Elie, Yannis Michalakis, Mircea T Sofonea, Antonin Bal, Sylvie Behillil, Grégory Destras, David Boutolleau, Sonia Burrel, Anne-Geneviève Marcelin, Jean-Christophe Plantier, Vincent Thibault, Etienne Simon-Loriere, Sylvie van der Werf, Bruno Lina, Laurence Josset, Vincent Enouf, Samuel Alizon, the COVID SMIT PSL group

## Abstract

France was one of the first countries to be reached by the COVID-19 pandemic. Here, we analyse 196 SARS-Cov-2 genomes collected between Jan 24 and Mar 24 2020, and perform a phylodynamics analysis. In particular, we analyse the doubling time, reproduction number (*ℛ*_t_) and infection duration associated with the epidemic wave that was detected in incidence data starting from Feb 27. Different models suggest a slowing down of the epidemic in Mar, which would be consistent with the implementation of the national lock-down on Mar 17. The inferred distributions for the effective infection duration and *ℛ_t_* are in line with those estimated from contact tracing data. Finally, based on the available sequence data, we estimate that the French epidemic wave originated between mid-Jan and early Feb. Overall, this analysis shows the potential to use sequence genomic data to inform public health decisions in an epidemic crisis context and calls for further analyses with denser sampling.

## Introduction

On Jan 8 2020, the Chinese Center for Disease Control announced that an outbreak of atypical pneumonia was caused by a novel coronavirus (Liu et al., 2020). The genetic sequence of what is now known as SARS-Cov-2 was released on Jan 10 (Wu et al., 2020; Zhou et al., 2020). This was less than two weeks after the initial report of the outbreak by the Wuhan Health Commission, which took place on Dec 31 2019. Never has a novel pathogen been sequenced so rapidly.

The number of sequences in the databases grew rapidly thanks to an altruistic and international effort of virology departments all around the world gathered via the Global Initiative on Sharing All Influenza Data (GISAID, https://www.gisaid.org/). Early results allowed better understanding the origin of SARS-Cov-2 and identification of a bat coronavirus (SARSr-CoV RaTG13) as its closest relative with more than 96% homology, as well as some potentially adaptive mutations (Andersen et al., 2020; ICTV, 2020; Xiao et al., 2020).

The available sequences were also analysed using the field of phylodynamics (Frost et al., 2015; Grenfell et al., 2004; EM Volz et al., 2013), which aims at inferring epidemiological processes from sequence data with known sampling dates. Most of these analyses were shared through the website virological.org. In particular, using 176 genomes from which he extracted 85 representative sequences (to avoid a potential cluster effect), (Rambaut, 2020) estimated the molecular clock to be approximately 8 · 10^−4^ substitutions per position per year, with a 95% Highest Posterior Density (HPD) between 1.4 · 10^−4^ and 1.3 · 10^−3^ subst./pos./year, which yielded a date of origin of the outbreak mid-Nov 2019, with a 95% HPD spanning from Aug 27 to Dec 19. Further analysis with more recent sequences found a median estimate of 1.1 • 10^-3^ subst./pos./year with asimilarHPD (Duchene et al., 2020). In their work, (Scire et al., 2020) explored a variety of priors for the analysis and found similar orders of magnitude for the molecular clock estimate. They also applied a birth-death model to estimate several parameters including the temporal reproduction number (*ℛ_t_*) but a difficulty is that not all sequences originated from China and the sampling rate could also vary. Finally, (E Volz et al., 2020) performed one of the early analyses of the outbreak using coalescent models, allowing them to estimate the date of the origin of the epidemic in early Dec 2019 (with a 95% CI: between 6 Nov and 13 Dec 2019) and the doubling time of the epidemic to be 7.1 days (with a 95% CI: 3.0-20.5 days). These reports mention several caveats, which are due to the limited number of sequences, the limited amount of phylogenetic signal, the potentially unknown variations in sampling rates and the sampling across multiple countries.

The first COVID-19 cases were detected in France from Jan 24, 2020, mostly from travellers, but these remained isolated until Feb 27, when the national incidence curve of new COVID-19 cases started to increase steadily. Limited measures were announced on Feb 28, but schools were closed from Mar 16, and a nationwide lock-down was implemented from Mar 17. On Apr 19, the prime minister gave the first official estimate of the basic reproduction number (*ℛ*_0_), which was 3.5, and of the temporal reproduction number after the lock-down, which was 0.5 (Salje et al., 2020).

We study the COVID-19 epidemic in France by analysing 196 genomes sequenced from patients diagnosed in France that were available on Apr 4, 2020 thanks to the GISAID and to French laboratories (see the online Supplementary Table for the full list). (Gambaro et al., 2020) provided a first picture of the general genomic structure of French epidemic using 97 genomes from samples collected in the north of France between Jan 24 and Mar 24, 2020. They identified several independent introductions of the virus in France but also found that the majority of the sequences belong to a major clade. This clade belongs to a larger clade labelled as G by GISAID, A2 by the nextstrain (http://nextstrain.org/) platform and B.1 following the dynamical taxonomy introduced by (Rambaut, Holmes, et al., 2020). We refer to it as the clade related to the epidemic wave.

Our early phylodynamics analyses focus on the epidemic doubling time, the generation time, and the temporal reproduction number *ℛ*(*t*). Current data does not allow us to perform a phylogeographic study and future work will investigate the structure of the epidemic within France, as well as potential dispersion between regions.

## Materials and Methods

### Data and quality check

On Apr 4,196 sequences were available from samples originating from France via the Global Initiative on Sharing All Influenza Data (GISAID, https://www.gisaid.org/) thanks to the work of the two Centre National de Référence and local virology laboratories. These sequences only provide a partial view of the epidemic as they originate from 8 the 18 French regions (Figure S1). Sequences were aligned and cleaned using the Augur pipeline developed by nextstrain (Hadfield et al., 2018). One sequence was removed due to low quality. The list of the sequences used is shown in the Online Supplementary Table.

We screened the dataset with RDP4 (Martin et al., 2015) using default parameter values and did not detect any recombination events.

### Phylogenetic inference

We first performed a maximum likelihood inference ofthe phylogeny using SMS (Lefort et al., 2017) and PhyML (Guindon and Gascuel, 2003). The mutation model inferred by SMS was GTR and was used as in input in PhyML. Other PhyML parameters were default. The resulting phylogeny was time-scaled and rooted using the software LSD (To et al., 2015) using a constrained mode with the sampling dates and a molecular clock rate fixed to 8.8 · 10^−4^ substitutions/position/year (the tree is provided in a Newick format in the Online Supplementary Materials).

We then used Beast 1.8.3 (Drummond, Suchard, et al., 2012) to perform inference using a Bayesian approach. More specifically, we assumed an exponential coalescent for the population model (Drummond, Nicholls, et al., 2002). We used the default settings for the model, which correspond to a gamma distribution for the growth rate prior Γ(0.001,1000) and an inverse prior for the population size *1/x* (see Supplementary Methods S1).

We also used Beast 2.3 (Bouckaert et al., 2014) to estimate key parameters using the birth-death skyline (BDSKY) model (Kühnert et al., 2014; Stadler, Kühnert, et al., 2013). One of these parameters is the temporal reproduction number (*ℛ_t_*) and we here assume three periods in the epidemic (which means we estimate 3 values *ℛ*_1_, *ℛ*_2_ and *ℛ*_3_). Another parameter is the the recovery rate, i.e. the rate at which the infectiousness ends. The final key parameter is the sampling rate, the inverse of which corresponds to the average number of days until an infected person is sampled. The ratio between the sampling rate and the sum of the sampling and the recovery rates indicates the fraction of infections that are actually sampled. By sampled, we mean that the patient is identified and the virus population causing the infection is sequenced. Note that we assume sampled hosts are not infectious anymore. We considered multiple priors for the rate of end of the infectious period by setting a lognormal prior LogNorm(90,0.5) and a uniform prior Unif(5,350). We assumed a beta prior (1.0,1.0) for the sampling rate (see Supplementary Methods S1). As in previous models (Stadler, Kühnert, et al., 2013), we set the sampling rate to 0 before the first infected host is sampled (here on Feb 21, 2020).

For both analyses in Beast, we assumed a GTR mutation model, following the results of SMS. We also assumed a uniform prior *U*(0, 1) for the nucleotide frequencies and a lognormal prior for parameter *κ*, LogNorm(1,1.25).

Regarding the molecular clock, earlier studies have reported a limited amount of phylogenetic signal in the first sequences from the COVID-19 pandemic. Given that we here focus on a subset of these sequences, we chose to fix the value of the strict molecular clock to 8.8 · 10^−4^ substitutions/position/year, following the analysis by (Rambaut, 2020). In Appendix, we study the influence of this value on the results by setting it to a lower (4.4 · 10^−4^ subst./pos./year) or a higher (13.2 · 10^−4^ subst./pos./year) value. Finally, we also estimate this parameter assuming a strict molecular clock. The most recent estimates suggest that the intermediate and high value are the most realistic ones (Duchene et al., 2020).

### Data subsets

We analysed subsets of the whole data set. Our largest subset excluded 10 sequences that did not belong to the French epidemic wave clade and therefore contained 186 sequences. Figure S1 shows the sampling date and French region of origin for each sequence. In general, the proportion of infections from the French epidemic that are sampled is expected to be in the order of 0.01%.

Some sampling dates are over-represented in the dataset, which could bias the estimation of divergence times (Seo et al., 2002; Stadler, Kouyos, et al., 2012). To correct for this, we sampled 6 sequences for each of the days where more than 6 sequences were available. This was done 10 times to generate 10 datasets with 122 sequences (France122a to France122h).

To investigate temporal effects using the coalescent model, we created three other subsets of the France122a dataset: “France61-1” contains the 61 sequences sampled first (i.e. from Feb 21 to Mar 12), “France61-2” contains the 61 sequences sampled more recently (i.e. from Mar 12 to Mar 24), and “France81” contains the 81 sequences sampled first (i.e. from Feb 21 to Mar 17).

With the exponential coalescent model (denoted DT for “doubling time”), we analysed all subsets of data (France61-1, France61-2, France81, and all the 10 France122 datasets), whereas for the BDSKY model we show the main dataset (France186) and analyse the 10 subsets with 122 leaves in Appendix.

## Results

### Phylogeny and regional structure

Figure 1 shows the regional structure of the French epidemic. Sequences corresponding to black leaves were ignored in the subsequent analyses because they do not belong to the main clade. Most of these originate from travelers isolated upon arrival in France, which explains their under-representation in the ongoing epidemic wave.

**Figure 1.**
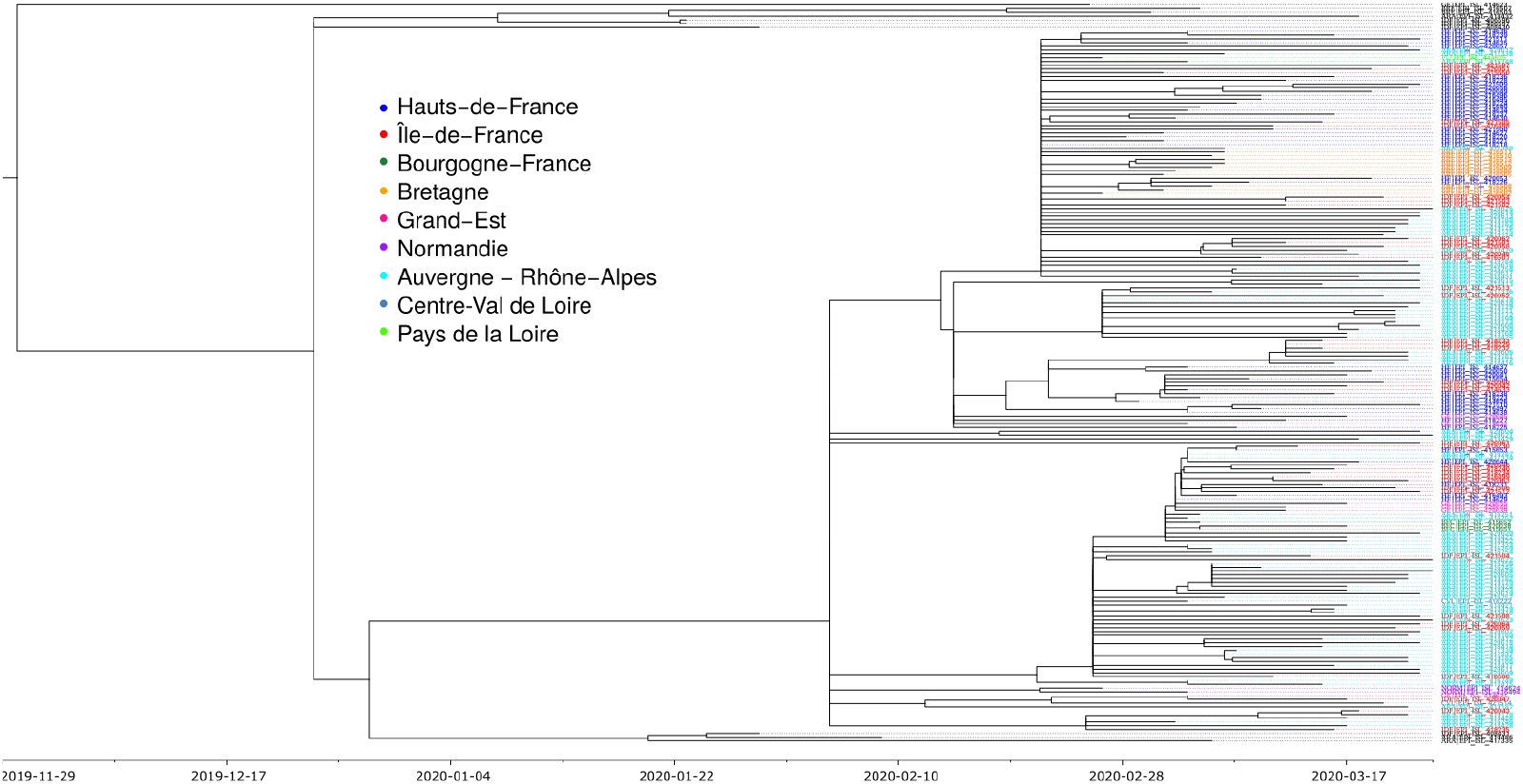
Phylogenetic structure of 196 SARS-Cov-2 genomes from France. Color shows the French region of sampling. Sequences in black were removed from the analysis because they fall outside the main clade corresponding to the epidemic wave. The phylogeny in a Newick format is available in the online Supplementary Materials.

Focusing on the main clade, we see that all the leaves originate from a common branching event, which is approximately half-point of the phylogeny. The polytomy in this point likely indicates a lack of phylogenetic signal. Addressing this issue will require more sequences from the early stages of the epidemic wave since currently the earliest sequence in this major clade is from Feb 21, 2020.

Colors indicate the regional structure of the French epidemic. As expected, we see some regional clusters. We also see that sequences from the same region belong to different subclades of the major clade, which is consistent with multiple introductions or dispersal between regions. Several French regions are not represented in the analysis. This is largely reflects the nature of the French COVID-19 epidemic, which has been stronger in the East of France and in the Paris area. This is also why this work focuses more on the speed of spread of the epidemic than on its general structure, which will be the focus of a future study (see also the work by (Gambaro et al., 2020)).

In the following, we focus on the main clade associated with the epidemic wave.

### Dating the epidemic wave

We first report the estimation of the time to the most recent common ancestor (TMRCA) of the 186 sequences that belong to the epidemic wave. Although this is the ancestor of the vast majority of the French sequences grouped in the B.1 clade (also referred to as G or A2 clade), the associated infection may have taken place outside France because the epidemic wave may be due to multiple introduction events (although from infections caused by similar viruses given the clustering).

Estimates of SARS-Cov-2 molecular clock should be treated with care given the limited amount of phylogenetic signal (Duchene et al., 2020; Rambaut, 2020). This is particularly true in our case since we are analysing a small subset of the data. In the Appendix, we present the analysis of the temporal signal in the data using the TempEst software (Rambaut, Lam, et al., 2016) and show that it strongly relies on early estimates that do not belong to the epidemic wave clade (Figure S2).

As shown in Figure 2, the molecular clock value directly affected the time to the most recent common ancestor for the coalescent model. This was also true for the BDSKY model, where the prior shape for the recovery rate, lognormal or uniform, had little impact compared to the assumption regarding the molecular clock (Figure S3). For both models, sampling of the 122 sequences amongst the 186 has a much smaller impact (Figure S5).

**Figure 2.**
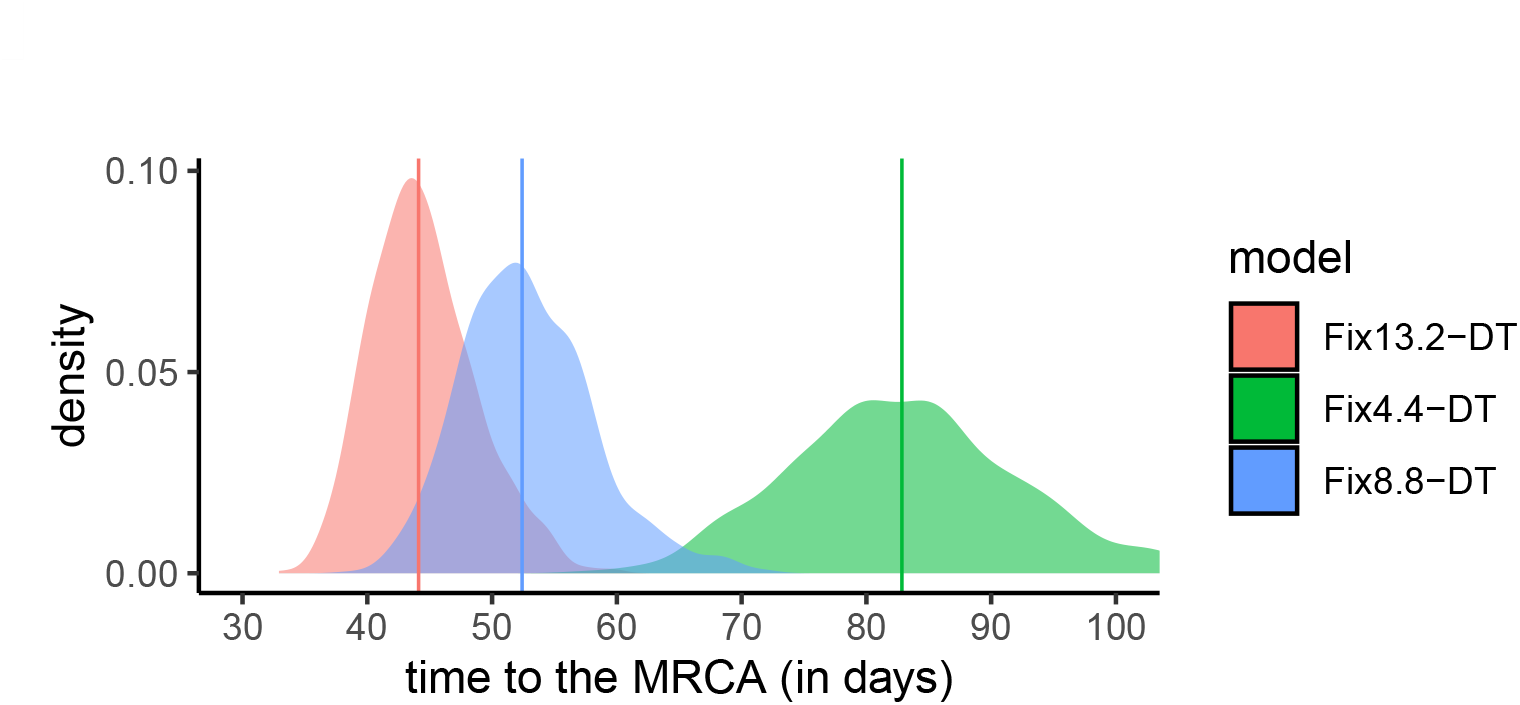
Time to the origin of the French epidemic wave as a function of the molecular clock. This estimate was obtained assuming an exponential growth coalescent population model and a fixed molecular clock (see Figure S4 for the BDSKY model). Colors indicate substitution rates and numbers (4.4, 8.8,13.4) refer values to be multiplied by 10^−4^ subsitutions/site/year. The slower the clock rate, the further away in time the most recent common ancestor (MRCA). Vertical lines show the distribution medians. The most recent sample dates from Mar 24, 2020.

Table 1 shows the dates for models with different evolution rates and different population models (exponential coalescent or BDSKY). Note that smaller datasets may not include the most recent samples.

**Table 1.**
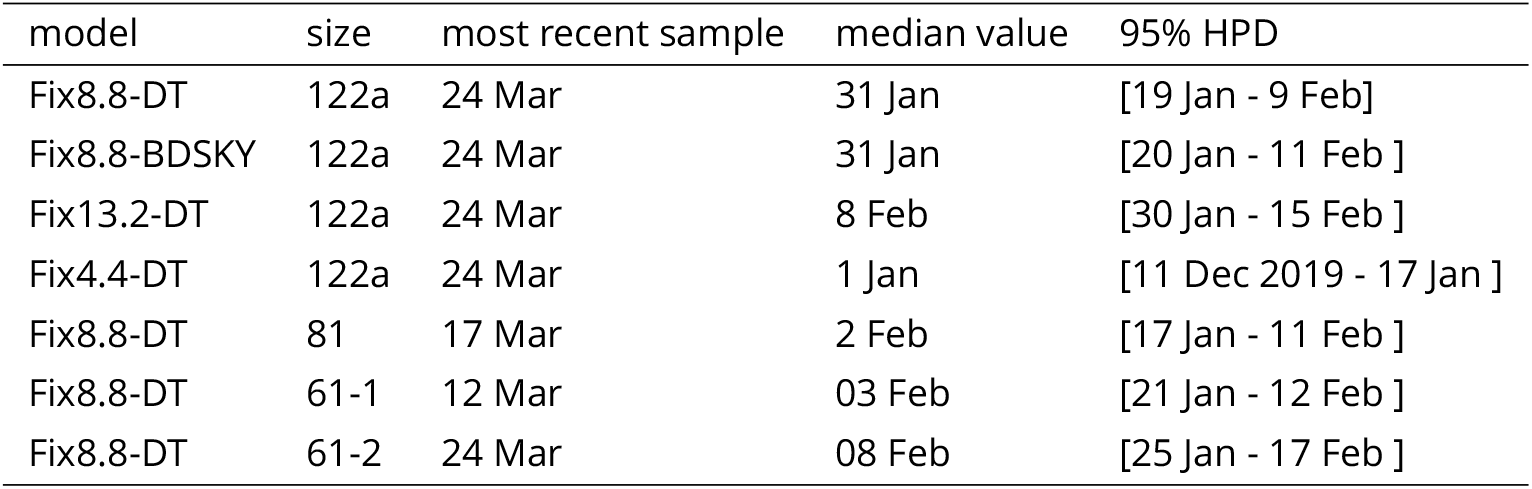
Date of the most recent common ancestor of the clade corresponding to the French epidemic wave. Unless specified otherwise, the year is 2020. The “model” indicates the value of the molecular clock and the population dynamics model used (DT or BDSKY).

For most of our datasets and models, the origin for the clade corresponding to the sequences from the French epidemic wave is dated between mid-Jan and early Feb. This large interval is due to the scarcity of “old” sequences (the first one collected in this clade dates from Feb 21) and on the fact that this clade averages the epidemic in several regions of France, which could have been seeded by independent introductions from outside France. The date provided by the slowest molecular clock (Fix4.4-DT) seems at odds with the data as we will see below.

To evaluate the effect of a potential sampling bias, we also estimate the time to the MRCA for 10 different sets of 122 sequences (Figure S5). We found similar median values for 9 of these 10 random datasets. Notice that the value of their parent dataset (France186), was slightly larger. For the BDSKY model, the effect was even less pronounced (Figure S4).

Overall, these dates (except for the slowest molecular clock) are consistent with those obtained by (Rambaut, 2020) regarding the beginning of the epidemic in China, which is dated November 17, 2019 with a confidence interval between Aug 27 and Dec 19, 2020. This interval is highly dependent on the number of available sequences as there are documented (but unsequenced) cases of COVID-19 in China early Dec 2019 (Li et al., 2020).

### Doubling time

Using a coalescent model with exponential growth and serial sampling (Drummond, Nicholls, et al., 2002), we can estimate the doubling time, which corresponds to the number of days for the epidemic wave to double in size. This parameter is key to calculate the basic reproduction number *ℛ*_0_ (Wallinga and Lipsitch, 2007).

In Figure 3, we show this doubling time for datasets that cover the whole (France122a), the first three quarters (France81), and the first half (France61-1) of the time period. Since the first dataset includes more recent sequences than the second, which itself includes more recent sequences than the third, our hypothesis is that we can detect variations in doubling time over the course of the epidemic. For completeness, we also show the results for the dataset covering only the second half of time period (France61-2).

**Figure 3.**
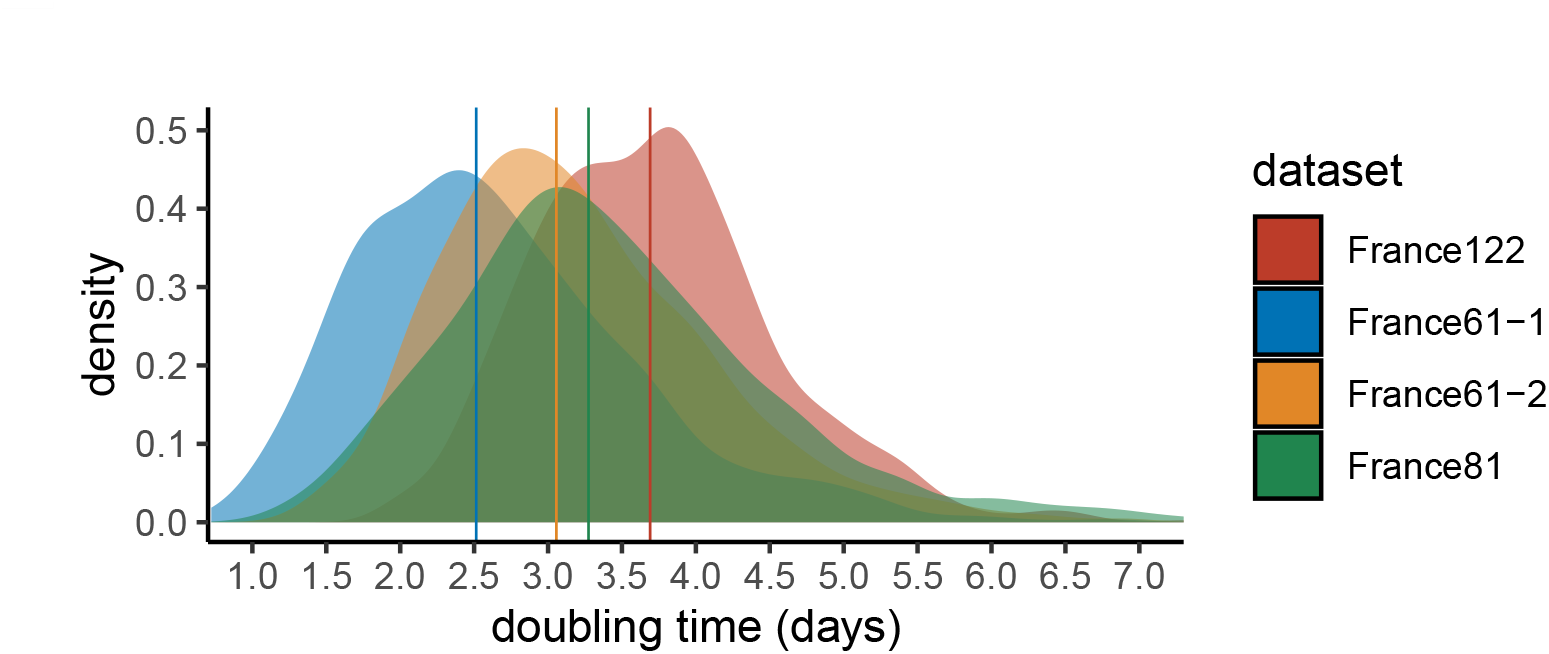
Epidemic doubling time. We assume an exponential growth coalescent model with a fixed molecular clock. The four datasets differ in the sequences analysed (see the Methods). Vertical lines show the distribution medians.

Adding more recent sequence data indeed leads to an increase in epidemic doubling time. Initially, with the first 61 sequences (which run from Feb 21 to Mar 12), the epidemic spreads rapidly, with a median doubling time of 2.5 days. With the addition of sequences sampled between Mar 12 and 17, the doubling time increases to 3.3 days. Finally, by adding sequences sampled between Mar 17 and 24, the doubling time rises to 3.7 days.

Importantly, the lower the number of sequences, the more the inferences become sensitive to the sampling scheme. This can be visualised with the fact that the doubling time obtained with the 61 most recent sequences (France61-2), which is, as expected, higher than that obtained using the 61 oldest sequences (France61-1), is lower than that obtained using all sequences (France122). Our interpretation is that phylogenetic signal becomes limited when only 61 sequences are considered. This can also be seen when estimating the date of origin of the epidemic: with the 61 most recent sequences, the date is comparable to that inferred using 122 sequences but assuming a faster evolutionary rate (Table 1). A more recent origin of the epidemic estimated with this subset of the data would directly lead to a lower epidemic doubling time.

To further explore the effect of sampling, we estimate the doubling time on 10 different sets of 122 sequences and find a limited effect on the median value (Figure S7). Notice that the value of the parent dataset (France186), is slightly larger.

We also study the effect of the molecular clock, i.e. the substitution rate, on the doubling time (Figure S6). As already mentioned above, the higher the molecular clock value, the lower the doubling time. However, for our realistic molecular clocks, the effect is limited: the median is 3.4 days assuming a high value for the molecular clock and 3.7 days for our default (medium) value. The low value of the molecular clock led to a high median doubling time of 5.6 days. This is at odds with the incidence data in France, which indicates an exponential growth rate of 0.23 days ^−1^ which corresponds to a doubling time of 3 days, suggesting that our default molecular clock is more realistic.

In comparison, phylodynamic inferences made from data from China with 86 genomes (Rambaut, 2020) found a median doubling time of about 7 days with a confidence interval between 4.7 and 16.3 days). One reason for the slower growth rate of the epidemic compared to ours is that we have focused on one rapidly expanding clade of the epidemic and neglected the smaller clades. Another possibility could be related to the timing of the sampling (early or late in the infection).

### Effective infection duration

The birth-death skyline (BDSKY) model (Stadler, Kuhnert, et al., 2013) allows us to estimate the effective duration of infection, which is defined in the model as the rate of becoming non-infectious (either through recovery, death, or sampling), and the reproduction number of the epidemic (i.e. the number of secondary infections caused by an infected host). The exponential growth coalescent model described above cannot distinguish between these two quantities. However, the BDSKY model requires more parameter values to be estimated.

The BDSKY model estimates separately the recovery rate and the sampling rate, and it is important to account for the latter because patients whose infections are sequenced can be assumed not to transmit the infection after this detection. The sampling rate after Feb 21 (it is set to 0 before that date) is estimated at 0.093 days ^−1^ with a (wide) 95% confidence interval between 0.006 and 0.627 days ^−1^. If we analyse this in days, the median value of the distribution yields 10.8 days and is consistent with the fact that in the French epidemic most of the screening for SARS-Cov-2 is done on severe cases upon hospital admission.

The distribution of infectious durations is obtained by taking the inverse of the sum of the sampling rate and the recovery rate. The median of this distribution is 5.12 days and 95% of its values are between 2.89 and 7.05 days (Figure 4). Note that this is an effective infection duration in that public health interventions can reduce it, e.g. by preventing transmission in the later stages of the infection, such that people can be infected but not infectious.

**Figure 4.**
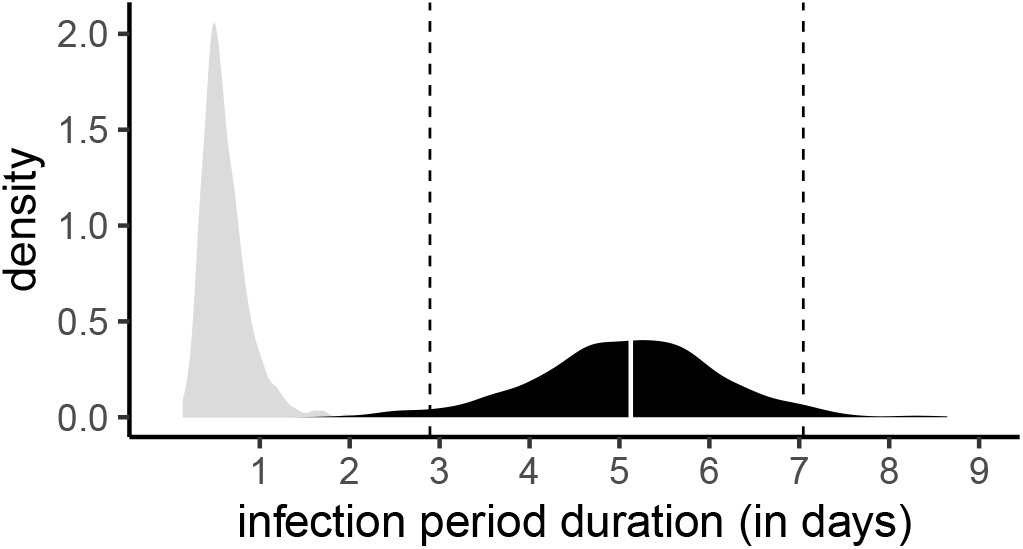
Distribution of effective infection duration. The prior distribution is shown in gray, and the posterior distribution in black. The white line shows the distribution median and the dashed line the 95% highest posterior density (HPD), which is between 3 and 7 days in agreement with results obtained using contact tracing data.

In Supplementary Figure S8, we show that the estimate for the effective infection duration is sensitive to the shape of the prior assumed for the recovery rate. Indeed, if we use a less informative (uniform) prior then the median sampling rate estimate is larger and the median infectious period estimated is shorter.

### Reproduction number

With the BDSKY model, we can estimate the temporal reproductive number, noted *ℛ*(*t*), since the onset of the epidemic wave. Here, given the limited temporal signal, we only divided the time into 3 intervals to estimate three reproduction numbers: *ℛ*_1_ before Feb 19, *ℛ*_2_ between Feb 19 and Mar 7, and *ℛ*_3_ between Mar 7 and Mar 24.

These results are very consistent with those obtained for the doubling time, even if the time periods are different. For the period before Feb 19, the estimate is the least accurate with values of *ℛ*_1_ with a median of 1.05 but at 95% Highest Posterior Density (HPD) between 0.13 and 3.22. The lack of information can be seen in Figure 5 as the posterior distribution (gray area) is very similar to the prior (dashed curve). This is consistent with the fact that the oldest sequence dates from Feb 21, while the tree root is estimated at the beginning of Feb. Over the second time period (in orange), the distribution shape is similar to that of the prior but the median is very different and rapid growth is detected with a median value of *ℛ*_2_ of 2.56 (95% HPD between 1.66 and 4.74). Finally, the most recent period after Mar 7 is the most accurate and detects a slowing down of the epidemic with a R_3_ of 1.38 (95% HPD between 1.13 and 2.03)

**Figure 5.**
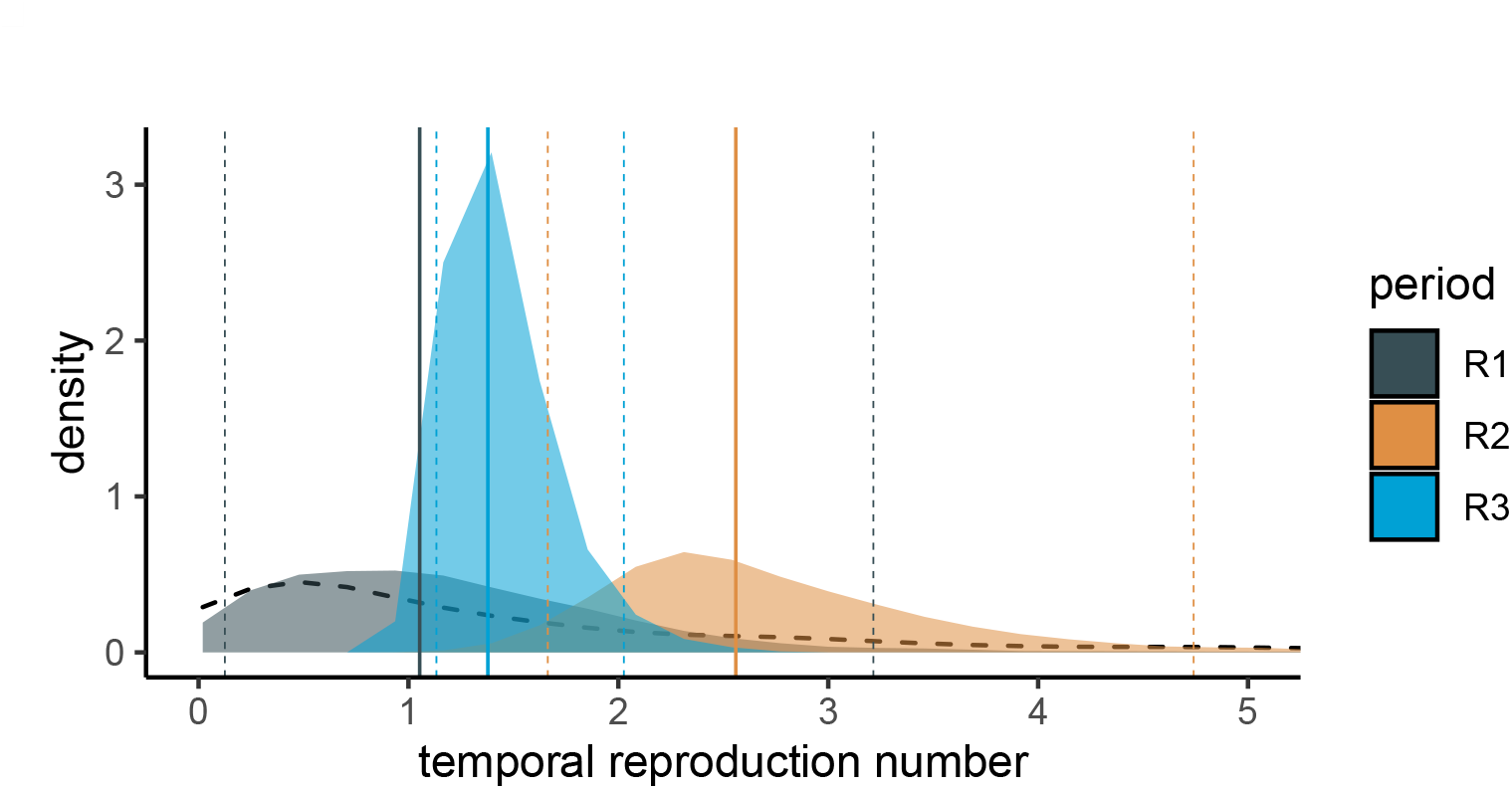
Temporal reproduction numbers inferred using the BDSKY model. These results are obtained for the France186 dataset. The black dashed curve show the prior distribution, and the posterior distributions are in color. Vertical plain lines show distribution medians, while vertical dashed lines indicate the 95% highest posterior density (HPD).

In Appendix, we show that these estimates for *ℛ_t_* are robust to the prior used for the recovery rate (Figure S9). They are also robust to the sampling of 122 of the 186 sequences (Figure S10).

## Discussion

Analysing SARS-Cov-2 genome sequences with a known date of sampling allows one to infer phylogenies of infections and to estimate the value of epidemiological parameters of interest (Frost etal., 2015; EM Volz et al., 2013). We performed this analysis based on the 196 sequences sampled in France and available on Apr 4, 2020. We focus in particular on the largest clade regrouping 186 of the most recent sequences and likely corresponding to the epidemic wave that peaked in France early Apr 2020.

Before summarizing the results, we prefer to point out several limitations of our analysis. First, the French clade we analysed is in fact an international clade: although most French sequences appear to be grouping into two main subclades within this clade, it is possible that the variations in epidemic growth that we detect are more due to European than French control policies. Second, some French regions (e.g. Auvergne-Rhone-Alpes) are more represented than others (e.g. Occitanie is absent), which could bias the analysis at the national level. However, the coverage is largely proportional to the state of the epidemics in France in March, where the Paris area and the East of France were more heavily impacted. Therefore, we expect the addition of sequences from less impacted regions to have a limited effect on our doubling time and reproduction number estimates. Finally, the molecular clock had to be set in this analysis because we do not have enough samples from the month of Feb in France.

Despite these limitations, our results obtained early Apr confirm a slowing down of the epidemic in France, where the epidemic peak in terms of ICU admissions was reached on Apr 1. Indeed, by adding sequences sampled between Mar 12 and 24 to the phylogeny, the doubling time of the epidemic estimated by an exponential growth coalescent model increased by 48%. This slowdown is more clearly detected using a birth death model via the temporal reproduction number *ℛ*(*t*): the median value decreased by 41% after Mar 12. This is consistent with the implementation of strict control measures in France as of Mar 17. These variations and even these orders of magnitude are consistent with our estimates based on the time series of incidence of new hospitalizations and deaths (Sofonea et al., 2020). However, these results were obtained with relatively few sequences and a denser sampling is needed to be more confident in our ability to detect an epidemic slowdown.

Finally, the BDSKY model also provides us with an estimate of the effective infection duration. This can be seen as the generation time of the epidemic, i.e. the number of days between two infections, and is an essential component in the calculation of *ℛ*_0_ (Wallinga and Lipsitch, 2007). The result we obtain, with a 95% Highest Posterior Distribution between 3 and 7 days and a median of 5.2, is highly relevant biologically and comparable to results obtained using contact tracing data. For instance, (Ferretti et al., 2020) estimated a serial interval, which corresponds to the time between the onset of the symptoms in a ‘donor’ host and that in a ‘recipient’ host, with a median of 5 days and a standard deviation of 1.9 days. To date, there is no estimate of the serial interval in France.

By increasing the number of SARS-CoV-2 genomic sequences from the French epidemic (and the number of people working on the subject), in particular sequences collected at the beginning of the epidemic, it would be possible to better estimate the date at which the epidemic wave took off in France, improve the estimate for the infection generation time and the reproduction number, better understand the spread between the different French regions, and estimate the number of virus introductions into the country.

Finally, it is important to set these results into their context. As acknowledged in the introduction, the French state only acknowledged the magnitude of the COVID-19 epidemic on the last days of Feb 2020 and these genomes were mostly collected between Feb 21 and Mar 24. Most of this analysis was published on Apr 6. At this time, the epidemic peak was barely noticeable in the incidence data. Furthermore, the serial interval, which is used to estimate the generation time of the infection and classically measured from contact tracing data, is still unknown in France. These results illustrate the contribution phylodynamics can make to public health during a crisis.

## Data Availability

Sequences data originated from the Global Initiative on Sharing All Influenza Data (GISAID, https://www.gisaid.org/) and are listed in Appendix Table.

https://platform.gisaid.org/

## Data accessibility

Data are available online at https://platform.gisaid.org/ with prior registration to the GISAID at www.gisaid.org

## Online supplementary material

A TSV file listing the sequences used and a text file containing the unrooted phylogeny in a Newick format can be found on www.medrxiv.org/content/10.1101/2020.06.03.20119925v2.supplementary-material

## Acknowledgements

We thank the patients, nurses, doctors, and all the French laboratories who made this work possible by generating and sharing the virus genome sequences. We also thank the ETE modelling group for discussion.

Gonché Danesh is supported by a doctoral grant from the Fondation pour la Recherche Médicale (FRM grant number ECO20170637560).

This work was partly supported by the *Urgence Recherche Covid-19* call from the Occitanie Region (contract 20007477) and the ANR (PhyEpi project).

We acknowledge the IRD itrop high-performance computer (South Green Platform) at IRD montpellier for providing resources that have contributed to the results presented in this work (more details on bioinfo.ird.fr).

Preliminary versions of this work were posted on Apr 9 (in French on http://covid-ete.ouvaton.org) and on Apr 21 (in English on http://virological.org/).

We thank Luca Ferretti and two anonymous reviewers for their carefull reading and numerous suggestions.

Version 3 of this preprint has been peer-reviewed and recommended by Peer Community In Evolutionary Biology (https://doi.org/10.24072/pci.evolbiol.100107)

## Cite as

Danesh G et al. (2020). Early phylodynamicsanalysis ofthe COVID-19 epidemic in France. *medRxiv* 2020.06.03.20119925, ver. 3 peer-reviewed and recommended by *PCI inEvolutionaryBiology*. doi: 10.1101/2020.06.03.20119925

## Posted

TBC

## Recommender

BJesse Shapiro

## Reviewers

Luca Ferretti and two anonymous reviewer

## Appendix

### S1 BEAST priors

MCMC chains were run for 5 · 10^8^ iterations. The first 10% runs were discarded as a burn in and convergence was assessed using Effective Sample Size (ESS). All parameters had ESS greater than 200.

Original XML files cannot be shared due to the GISAID agreement.

**Table S1.**
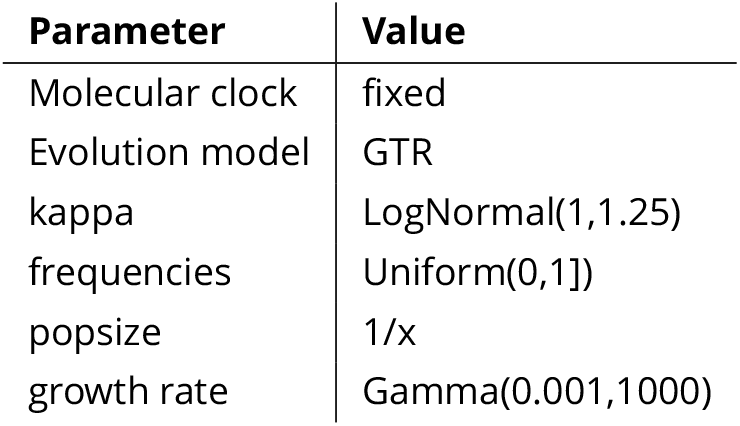
Prior summary for the exponential coalescent model

**Table S2.** Prior summary for the BDSKY model

| Parameter | Value |
| --- | --- |
| Molecular clock | fixed |
| Evolution model | GTR |
| kappa | LogNormal(1,1.25) |
| frequencies | Uniform[0,1] |
| Rate of end of infection | Uniform(1.2, ∞) or LogNormal(0,1.2) |
| Sampling rate | Beta(1,1) |
| Reproduction number | LogNormal(0,1.2) with maximum at 10 |

### S2 Supplementary figures

**Figure S1.**
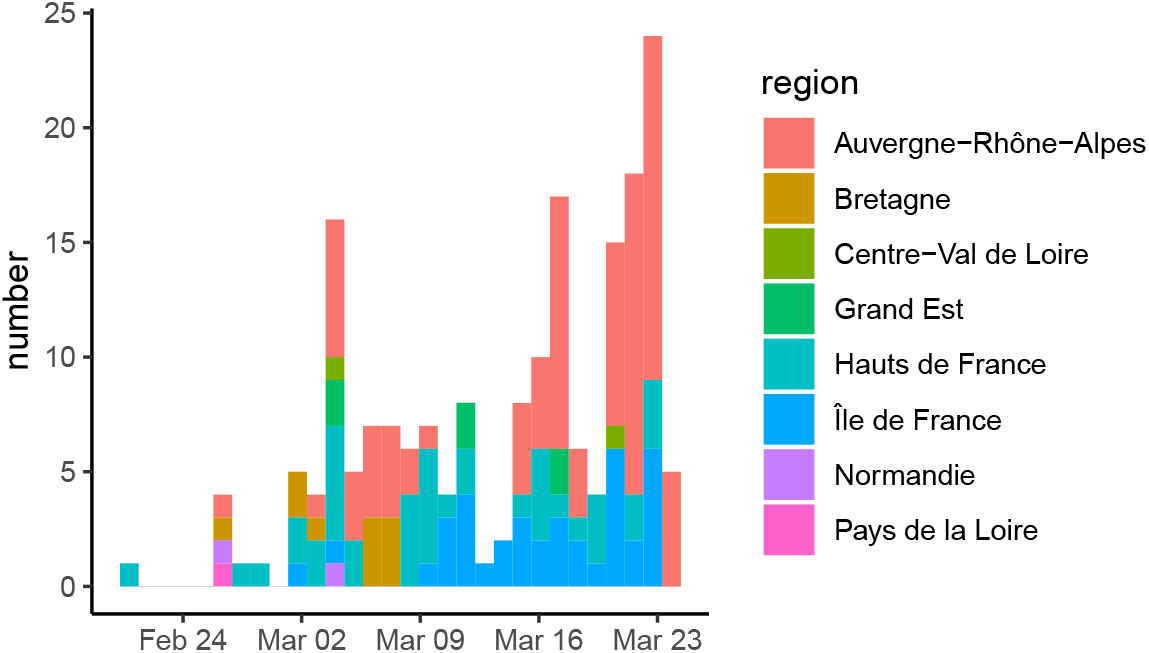
Sampling date and region. List of samples collected, analysed and shared via GISAID by the two French National Reference Centers (CNR) as of Apr 4, 2020.

**Figure S2.**
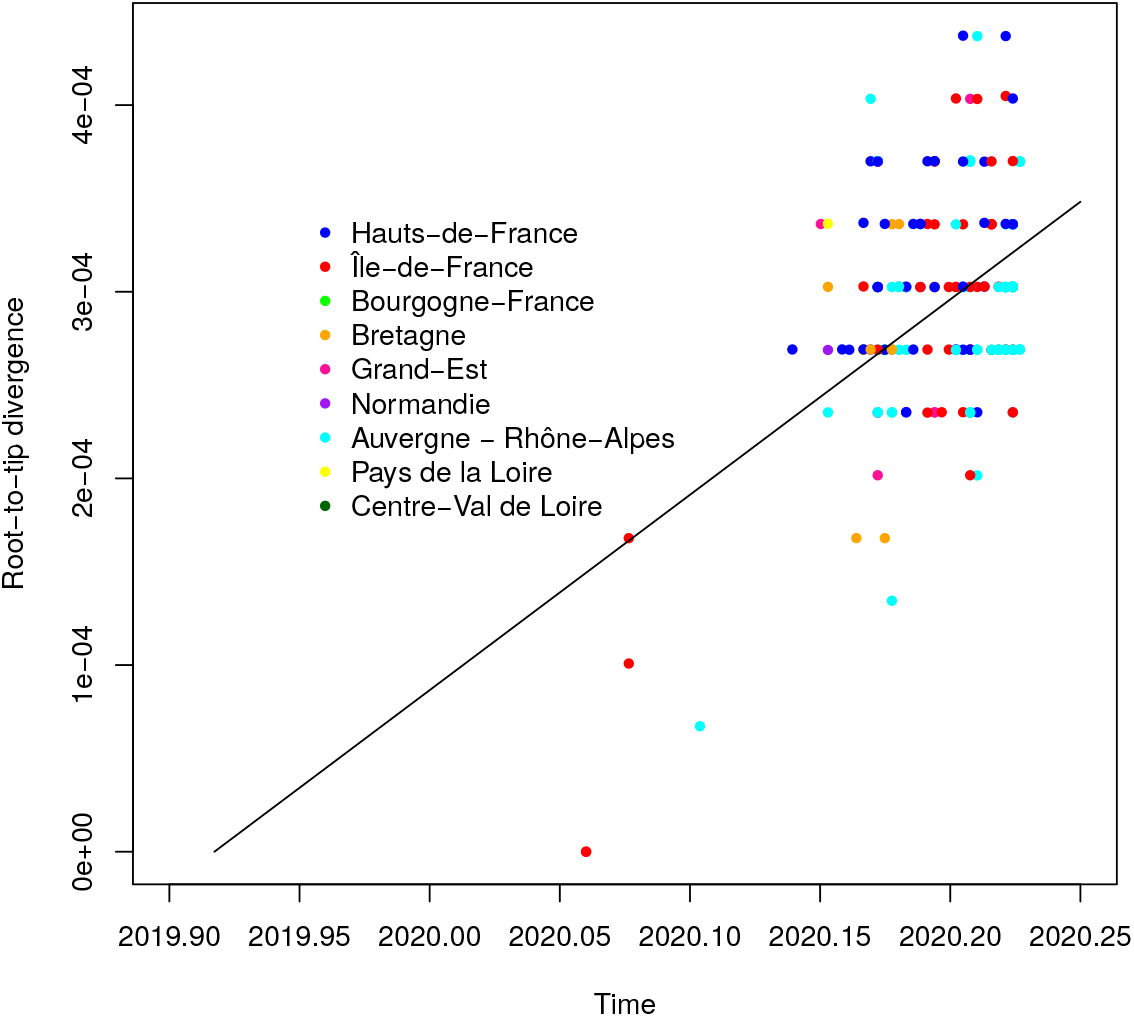
Root-to-tip correlation. We analyse a phylogeny based on all 196 French sequences (i.e. not only that from the epidemic wave). The four earliest cases in Jan and early Feb were all isolated and belong to another clade than the rest of the sequences. The figure was obtained using TempEst (Rambaut, Lam, et al., 2016).

**Figure S3.**
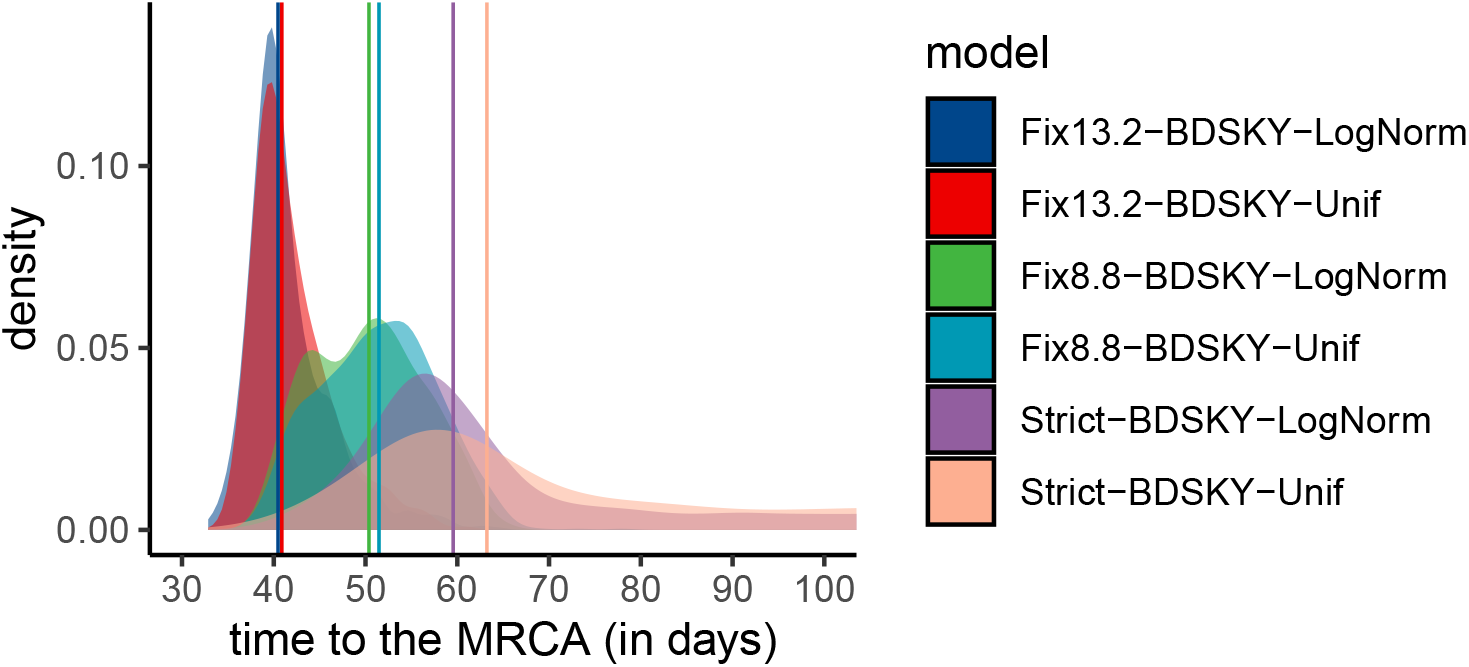
Time to the MRCA as a function of the molecular clock and of the recovery rate prior. Here we assume BDSKY model. Note that the posterior distributions are not very informative when the molecular clock value is estimated (the ‘Strict’ model).

**Figure S4.**
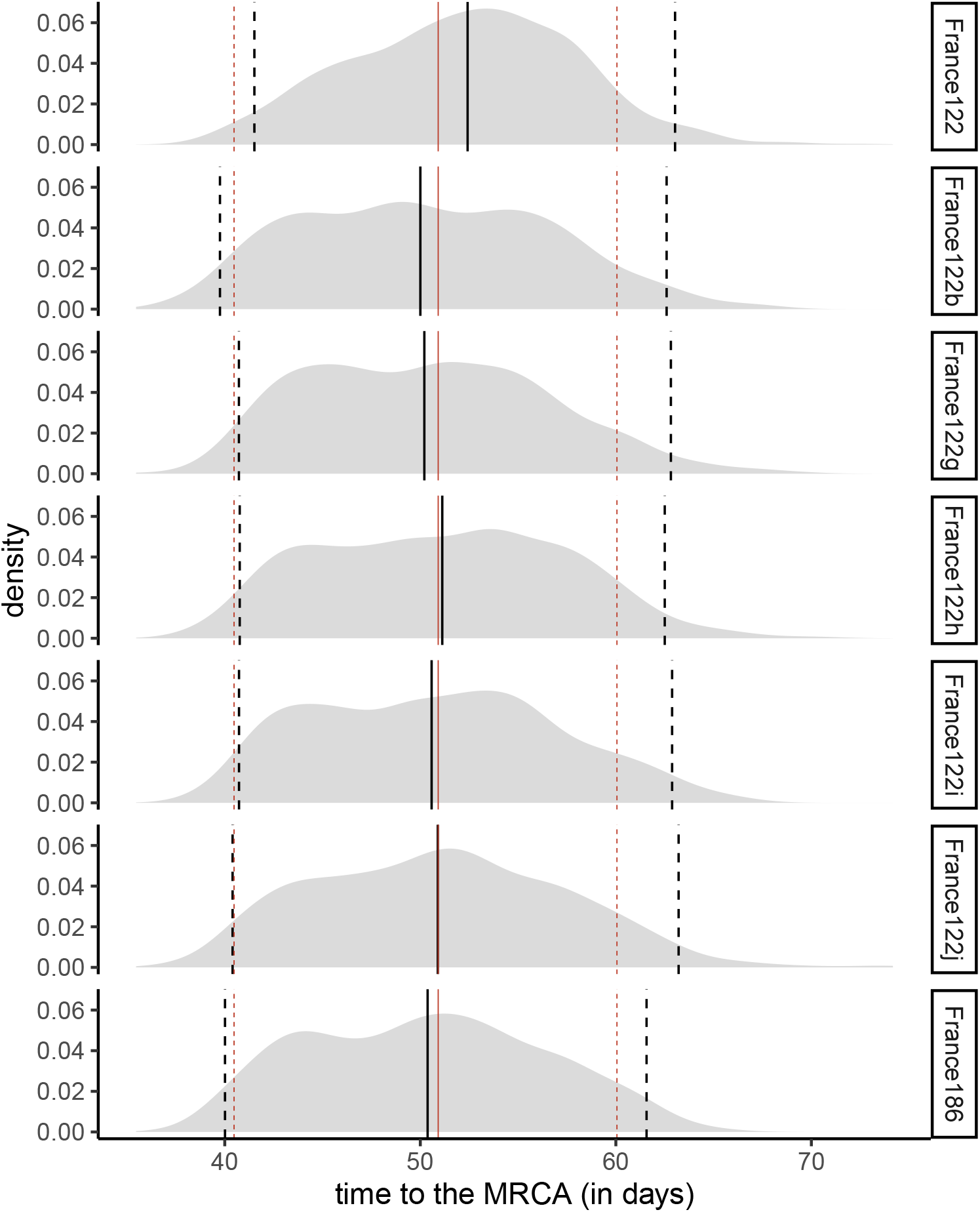
Time to the MRCA for the France186 dataset and subsets with 122 sampled sequences assuming the BDSKY model. The red lines show the quantiles (0.025, 0.5, and 0.975) for the average of the 10 datasets. The black line shows the quantile for each dataset, and we can see how it behaves compared to the average. The last panel shows the largest phylogeny built without sampling, i.e. using all 186 sequences.

**Figure S5.**
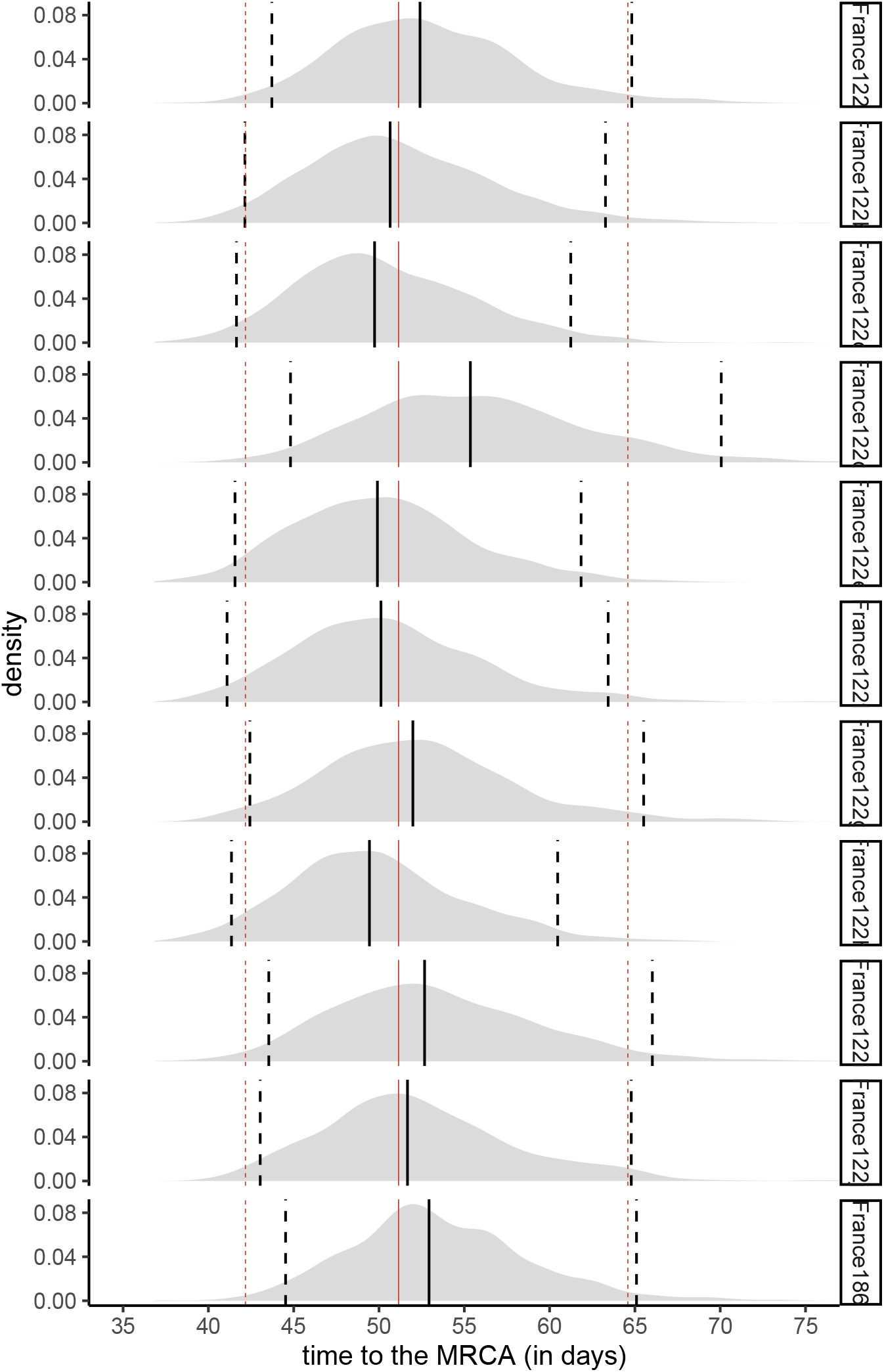
Time to the MRCA for the France186 dataset and subsets with 122 sampled sequences assuming an exponential growth coalescent model. The red lines show the quantiles (0.025, 0.5, and 0.975) for the average of the 10 datasets. The black line shows the quantile for each dataset, and we can see how it behaves compared to the average. The last panel shows the largest phylogeny built without sampling, i.e. using all 186 sequences. The latter phylogeny has a slightly larger number of days to the MRCA.

**Figure S6.**
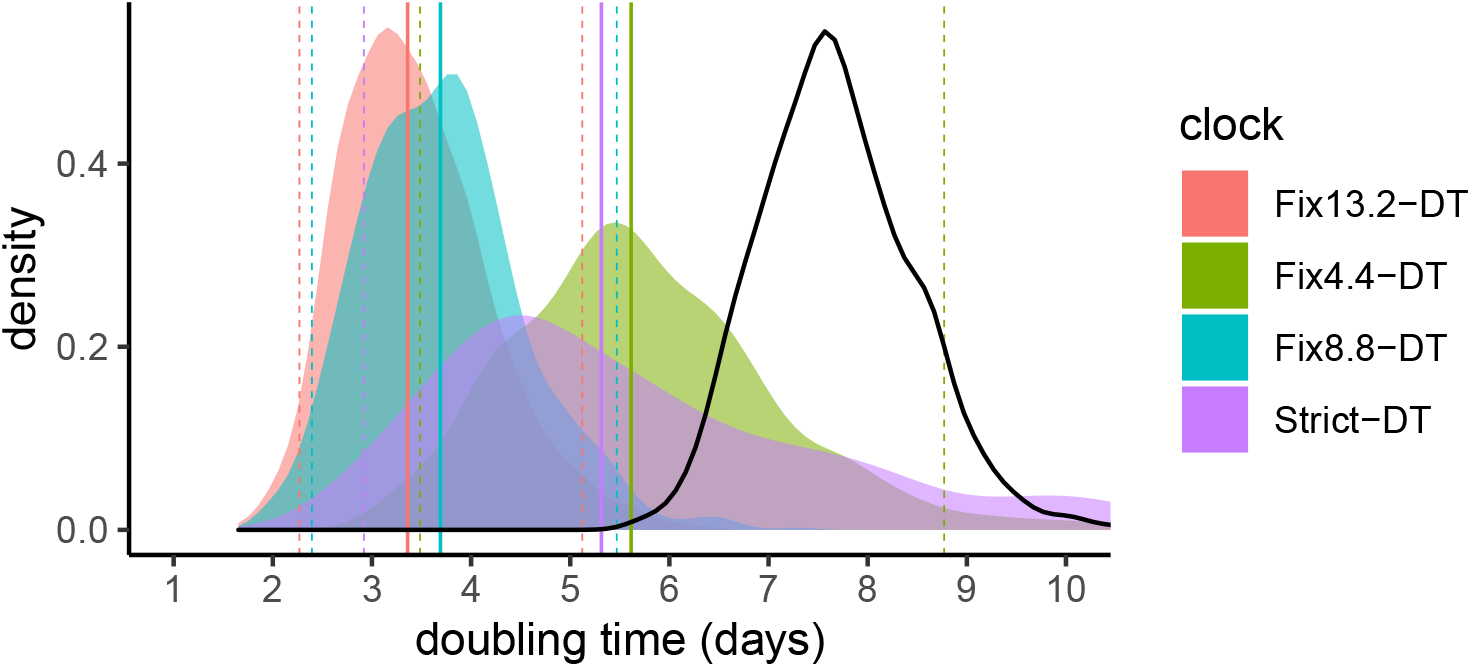
Effect of the molecular clock on the doubling time. Note that in the “Strict” model we infer the value using a strict molecular clock but the width of the posterior distribution is large which indicates a lack of phylogenetic signal. The thick black line shows the prior distribution (the true prior values do not allow to see the regular plot so we use here Gamma(3,100) instead of Gamma(0.001,1000)). Here, an exponential growth coalescent model is assumed and the dataset used is France122a.

**Figure S7.**
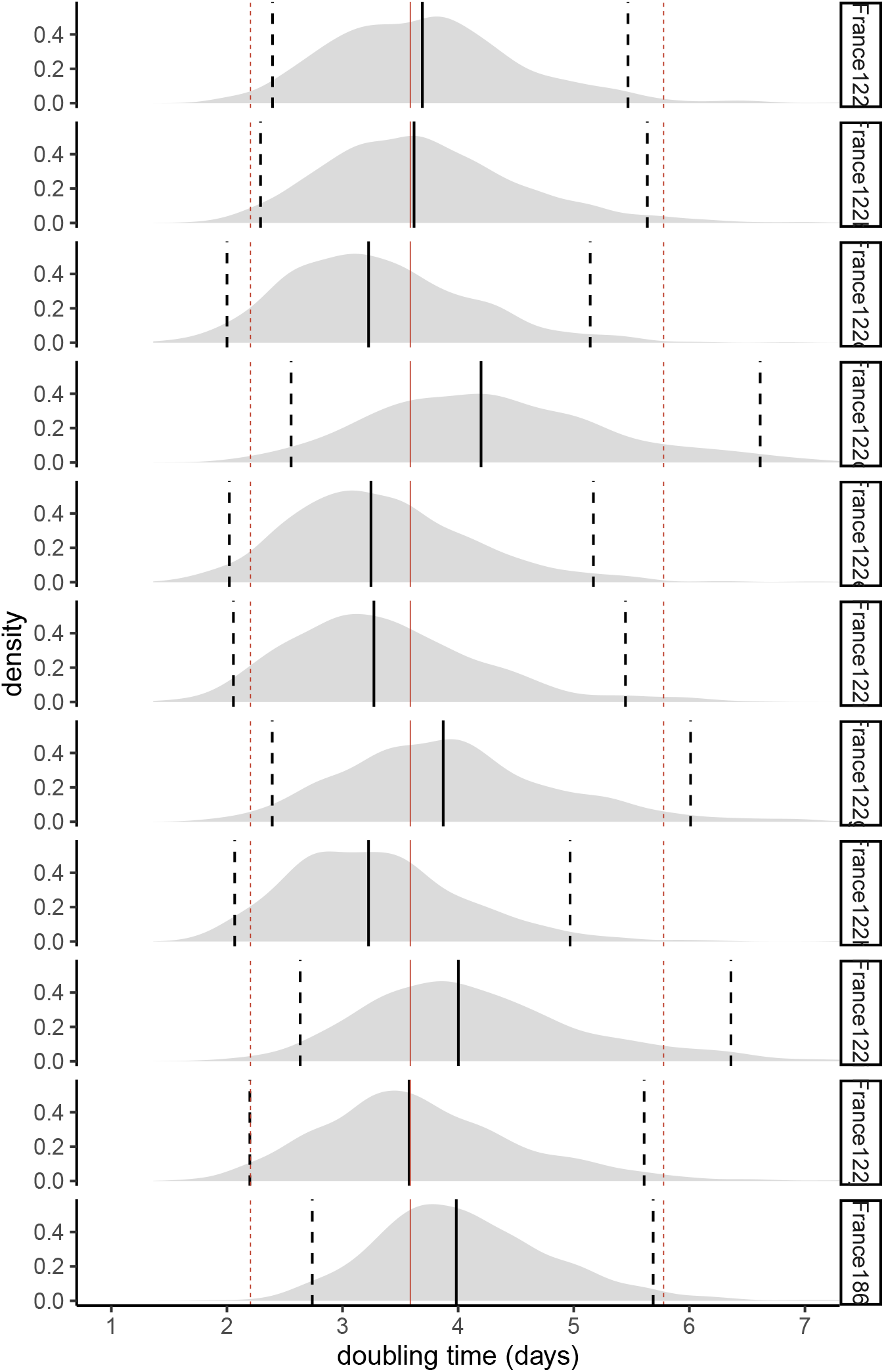
Doubling time for the 10 trees with 122 leaves sampled. The red lines show the quantiles (0.025, 0.5 and 0.975) for the average of the 10 datasets. The black line shows the quantile for each dataset. The last panel shows the largest phylogeny with 186 leaves (which slightly overestimates the doubling time).

**Figure S8.**
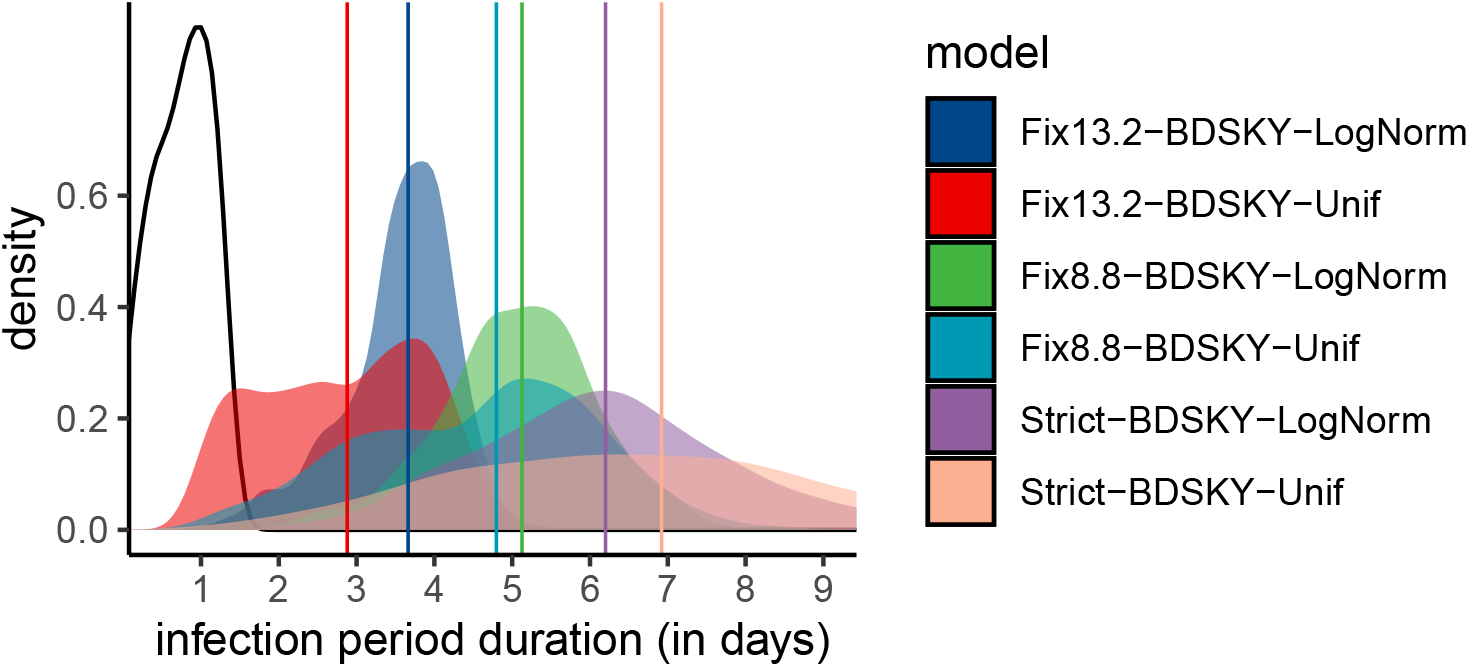
Effect of the prior shape and of the molecular clock on the effective infection duration estimate. The molecular clock value has a stronger effect than the prior shape. The thick black line shows the prior distribution. Note that the posterior distributions are not very informative, i.e. the 95% HPD is very large, when the molecular clock value is estimated (the ‘Strict’ model).

**Figure S9.**
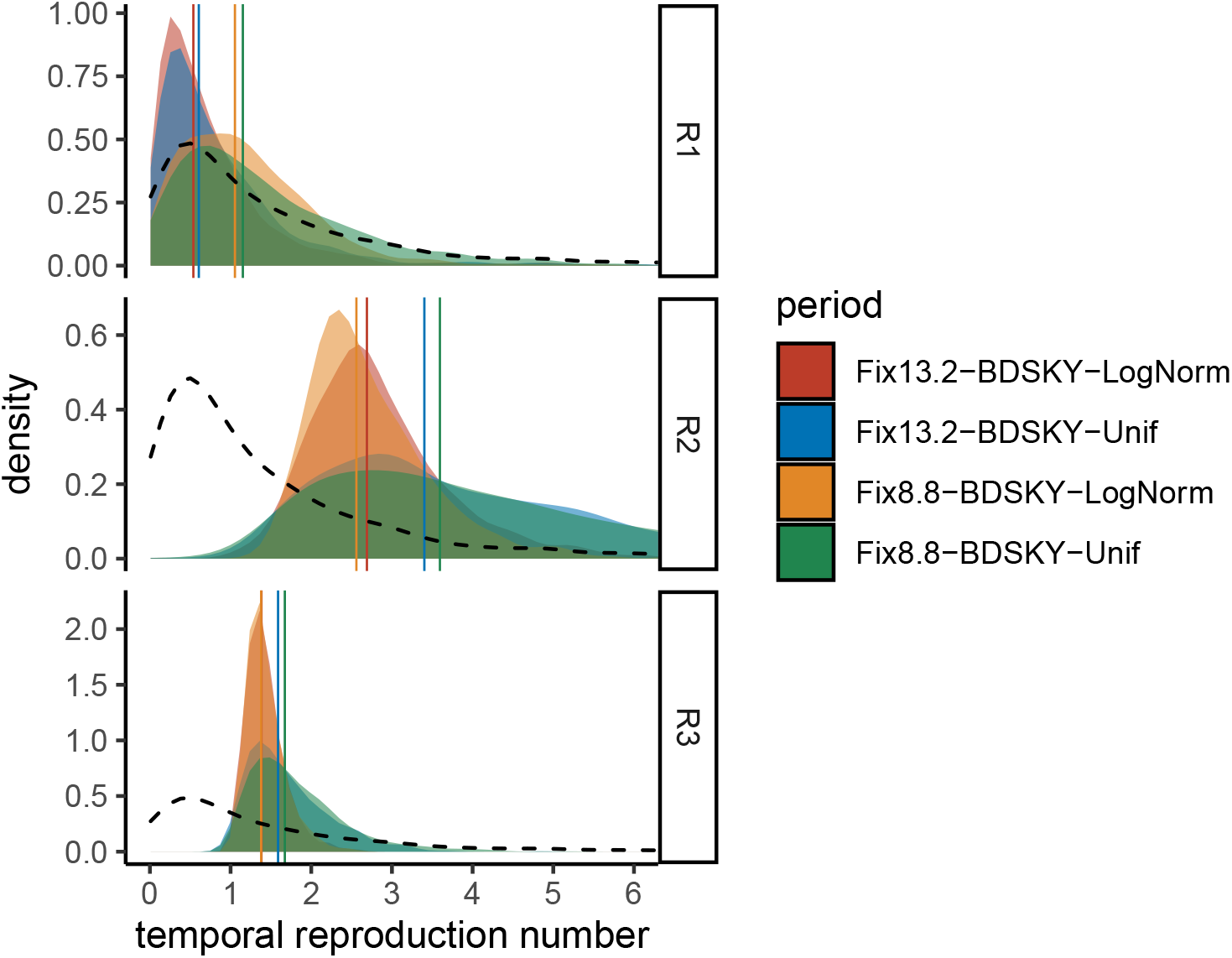
Effect of the prior shape and of the molecular clock on the temporal reproduction number estimate. The molecular clock value has a stronger effect than the prior shape except for *ℛ*_3_, where the effect is limited. For *ℛ*_1_, posterior distributions are close to the prior (black dashed line), as discussed in the main text.

**Figure S10.**
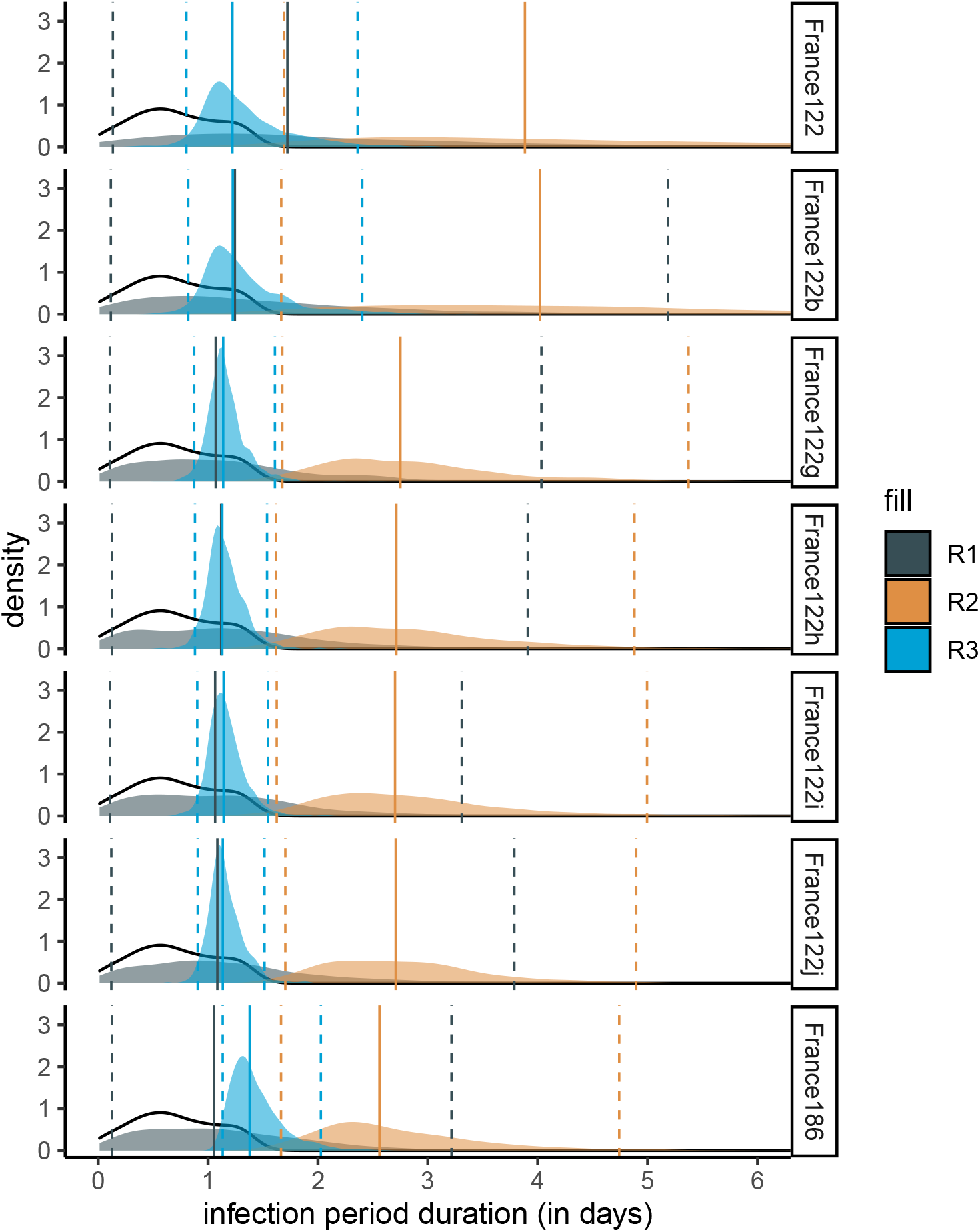
Reproduction numbers for the France186 dataset and subsets with 122 sequences. Here we assume BDSKY model. The thick line shows the prior distribution. For Ri, posterior distributions are close to the prior (black dashed line) indicated limited inference power.

**Figure S11.**
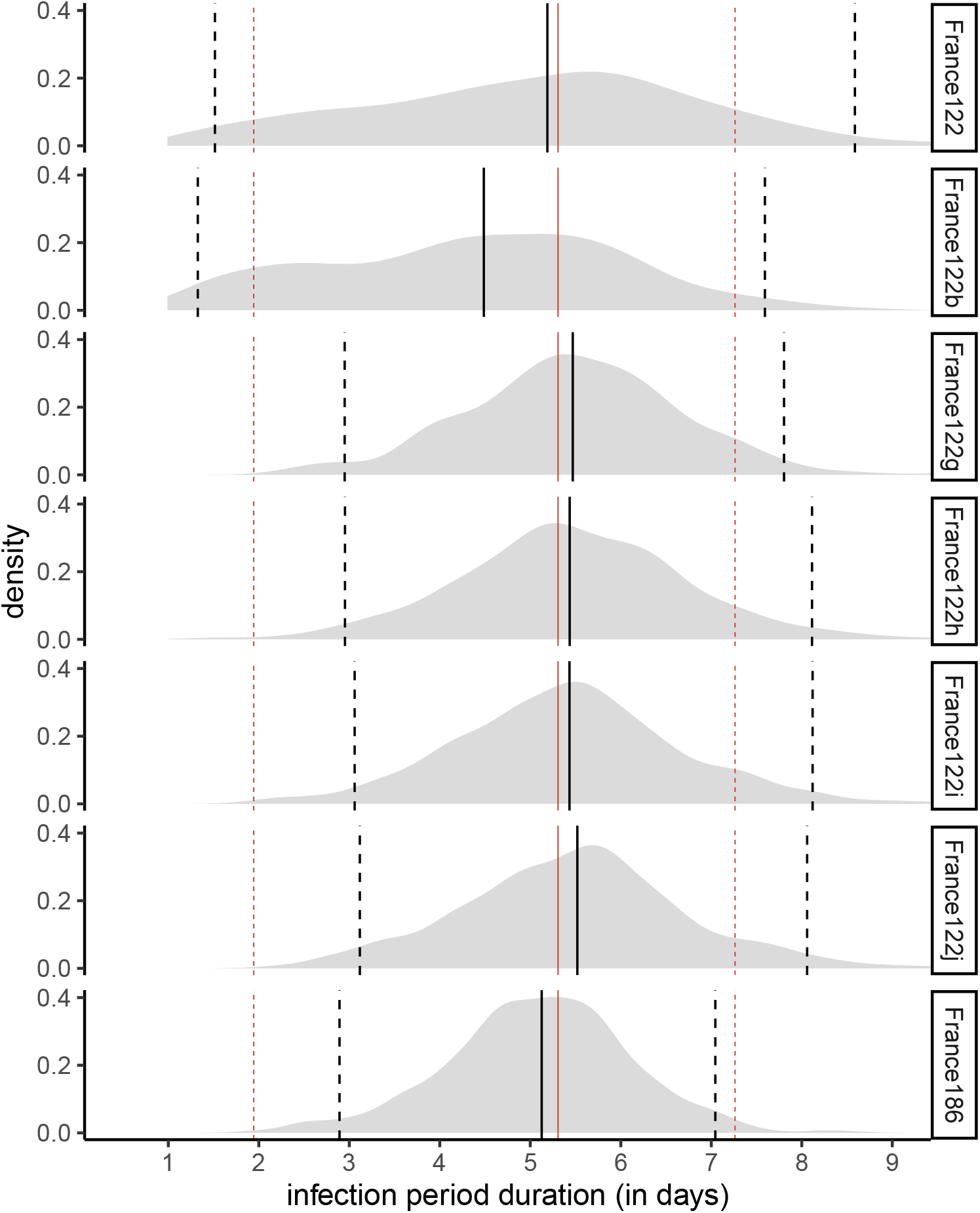
Effective infection duration for the France186 dataset and subsets with 122 sequences assuming a BDSKY model. The median value obtained with the whole dataset (France186, black full line) is close to the average of the median values obtained with the subsets (red full lines).

